# Socioeconomic disparities in substance use disorder prevalence and severity across the life course

**DOI:** 10.64898/2026.09.03.26362182

**Authors:** Peter B. Barr, Megan E. Cooke, Sally I-Chun Kuo, Henri M. Garrison-Desany, Gayathri Pandey, Kathleen K. Bucholz, Howard J. Edenberg, Sivan Kinreich, Bernice Porjesz, Jessica E. Salvatore, Jacquelyn L. Meyers

**Author notes:** Corresponding author: Peter B. Barr, Department of Psychiatry and Behavioral Sciences SUNY Downstate Health Sciences University, 450 Clarkson Ave, MSC 1203, Brooklyn, NY 11203.

## Abstract

Gradients across socioeconomic status (SES) exist across most health conditions. While there has been significant attention devoted to the relationship between SES and physical and mental health, less research has examined how these relate to substance use disorders (SUD). In the current study, we use data from the prospective study within the Collaborative Study on the Genetics of Alcoholism (COGA) to explore the relationship between SES (in early life and adulthood) with multiple substance use disorders (alcohol, tobacco, cannabis, opioid, cocaine, and other substances). We find that early life SES (parental education and income) are associated with lower odds of lifetime diagnoses and lower severity for tobacco (TUD) and cannabis (CUD) use disorders. Associations with TUD remain after adjusting for demographic and familial risk, but associations with CUD were null after including familial risk factors. In participants aged 25+, adult education was associated with lower odds of diagnosis and severity for each of the SUD considered, conditional on early life SES, adult income, sociodemographic characteristics, and familial risk for SUD. Exploratory analyses of changes in SES revealed that those who were upwardly or downwardly mobile were at the lowest or greatest risk for more severe SUD across multiple substances, respectively. Our results demonstrate the relevance of both early life and adult SES in SUD risk. Early life conditions seem particularly relevant for specific substances while greater adult education was associated with reduced risk across all forms of SUDs.

## Introduction

The gradient in health across socioeconomic status (SES) is one of the most well-documented phenomena in social epidemiology (Braveman et al., 2011). While significant attention has been devoted to understanding the relationship between SES and physical and mental health broadly, less research has systematically examined the relationship between SES and substance use disorders (SUD), especially beyond alcohol and tobacco use disorders.. Given the rapidly evolving landscape surrounding cannabis legalization (Cerdá et al., 2020), the increase in opioid-related deaths (Wilson et al., 2020), and overall decline in life expectancy due to substance-related disorders (Case and Deaton, 2015; Tilstra et al., 2021), identifying risk factors shared across various SUDs is vital for improving public health. Disentangling the etiologic pathways to each disorder, both within and outside the context of co-occurring SUDs, can help in identifying intervening mechanisms that may impact SUD. Previous research has pointed to the multifactorial risk for SUD, including adverse social conditions, adverse life events, neurodevelopmental, and genomic risk factors. The current research affords the opportunity for a more accurate and nuanced understanding of how these disorders occur, to aid prevention efforts of this pressing public health issue.

Socioeconomic status (SES), typically measured as a combination of income, education, and occupational status, is associated with a plethora of health outcomes (Adler and Ostrove, 1999; Braveman et al., 2011). Individuals at the lower end of the SES continuum are at higher risk for a range of adverse physical and mental health conditions, including SUDs (Adler and Rehkopf, 2008; Pampel et al., 2010). Theoretical mechanisms linking SES and poorer health under the “social causation” model include increased exposure to stressful and adverse conditions (e.g. financial strain, food insecurity, substandard living conditions) (Lloyd and Turner, 2008; Turner and Avison, 2003; Turner and Lloyd, 2003) and fewer resources to cope with adverse conditions (e.g., instrumental support, social networks, self-efficacy) (Pearlin et al.,

1981; Pearlin and Schooler, 1978). This increased exposure to chronic strains and limited coping resources embeds itself biologically through increased levels of inflammation (Muscatell et al., 2020), accelerated biological aging (Schrempft et al., 2022; Simons et al., 2022; Steptoe and Zaninotto, 2020), and poor health behaviors, such as smoking, lack of exercise, and unhealthy diets (Pampel et al., 2010), among others. Under the “social selection” model, poor early life health can impair one’s ability to maintain employment/educational opportunities, limiting future socioeconomic mobility (Blane et al., 1993; Haas, 2006). Thus, potential mechanisms linking SES and health are complex and multifactorial (Link and Phelan, 1995) and require large-scale studies with comprehensive measures to investigate.

A number of studies have documented associations between domains of SES, substance use, and problematic use (e.g., heavy use, SUDs). Adolescent tobacco, alcohol, and cannabis use are all associated with increased odds of high school dropout (Townsend et al., 2007), while higher educational attainment is associated with lower rates of SUD diagnoses by adulthood (Breslau et al., 2008; Kessler et al., 1995; Stinson et al., 2006). Associations between SUDs and education appear to be reciprocal, with evidence supporting both the social causation and social selection models (Breslau et al., 2011, 2008; Fothergill et al., 2008; Green et al., 2012; Martin et al., 2015; Verweij et al., 2013). Unemployment is associated with higher rates of SUD (Nolte-Troha et al., 2023), reflecting bidirectional associations between substance use behaviors and labor market participation over time (Boden et al., 2017; Jørgensen et al., 2019). According to U.S. national data (McKetta et al., 2021) rates of binge drinking are highest among those in lower prestige occupations (e.g., construction and extraction, sales, and installation, maintenance, and repair occupations), though in recent years the rate of binge drinking has increased more among those in high prestige occupations (e.g., legal, architecture and engineering, arts, design, entertainment, sports, and media occupations). Likewise, in a study of death certificates from Massachusetts (2011-2015) opioid-related overdose deaths were highest among those with physically demanding occupations; such as construction and extraction, farming, fishing, and forestry, health care support, and food preparation and serving (Hawkins et al., 2019; McKetta et al., 2021). Analyses of the National Epidemiologic Survey on Alcohol and Related Conditions III data reveal that adults with an annual family income of less than $20,000 were 50% more likely to meet criteria for a severe alcohol use disorder and 120% more likely to meet criteria for a severe drug use disorder compared to those with an annual income of $70,000 or more (Grant et al., 2016, 2015). In total, the evidence supports bidirectional associations between SES with SUD.

Despite evidence linking SES and SUD, relatively few studies have taken a life course perspective to examine the prospective associations between childhood SES and subsequent risk of SUD in adolescence and adulthood. Given prior work on the “long arm” of childhood SES and health (Hayward & Gorman, 2004), and the fact that by the end of high school, many adolescents report having used alcohol, marijuana, or nicotine (24-61.5%) and a notable minority report having used cocaine and opioids other than heroin (∼4-5%) (Miech et al., 2026), understanding the impact of early life conditions is critical for future risk of SUD. Kendler and colleagues (2014) found that parental SES during childhood (income, education, occupation) was positively associated with frequent alcohol use and negatively associated with alcohol problems across adolescence. Similarly, Barr and colleagues (2018) found that childhood SES was associated with trajectories of alcohol problems into young adulthood, but the direction of association varied across life course epochs, such that low SES was associated with increasing risk in adulthood. Although many young adults “mature out” of risky substance use behaviors (Lee and Sher, 2018; O’Malley, 2004), for others this period is marked by the onset of SUDs (Cooke et al., 2026). Individuals under the age of 35 account for 34% of opioid overdose deaths (Murphy et al., 2024; Wilson et al., 2020). Yet, the majority of longitudinal studies examining the development of substance use and problems focus on adolescence and end in the early 20s, missing later life stages. As a result, parental SES is often entangled with offspring SES, despite substantial SES shifts that can occur after young adulthood. Accordingly, the interplay between socioeconomic factors and the development of SUD are not well-characterized beyond the early 20s, despite SUD onsets being quite common well into the early 30s (Capaldi et al., 2015; Schulenberg et al., 2015).

SUDs also frequently co-occur, with comorbidity being the norm, which has led to the theory that the risk is best explained by a broad underlying genetic liability to addiction. There is substantial evidence from both the twin literature (Kendler and Myers, 2014; Krueger et al., 2007) and modern genome wide analyses (Hatoum et al., 2021; Karlsson Linnér et al., 2021; Poore et al., 2026) to support an underlying dimension of genetic risk across substances. Therefore, substance-specific disorders are not best studied in isolation when trying to understand downstream consequences of broad risk factors, yet few study samples have the capacity to examine multiple SUDs at once, and most studies focus primarily on alcohol use disorder. SUDs, beyond alcohol and tobacco occur at relatively lower rates in the broader population, with prevalence of 12-month and lifetime SUD at 3.9% and 9.9%, respectively (Grant et al., 2016) and rates appear to be declining in recent birth cohorts (Grucza et al., 2017). Therefore, samples enriched for substance use and use disorders, such as the Collaborative Study on the Genetics of Alcoholism (COGA), are needed to fully understand this transdiagnostic risk.

In the current study, we build on existing work in several key ways. First, drawing on data from the prospective study within COGA, a high-risk family study, we leverage the SUD-enriched nature of this sample to simultaneously explore multiple forms of substance use disorder (alcohol, tobacco, cannabis, opioid, cocaine, and other substances). Second, using data spanning childhood into early midlife, we investigate a large period of the life course which represents the most critical timing for SUD onset. Third, we explore the role of SES in both early life (e.g. parental SES) and in adulthood for SUDs to understand whether early life conditions have independent associations beyond current levels of attainment. Finally, given that COGA is a family-based design, we incorporate measures capturing familial and genetic risk, an important consideration in the etiology of substance use disorders.

## Methods

### Sample

The Collaborative Study on the Genetics of Alcoholism (COGA), ascertained high-risk families through adult probands in treatment for alcohol dependence and community-ascertained comparison families (Agrawal et al., 2023). Probands and all willing first-degree relatives were assessed; recruitment was extended to include additional relatives in families that contained 2 or more first degree relatives with alcohol dependence (n = 16,848). The COGA sample is 60.6% non-Hispanic White, 24.9% African American, 11.1% Hispanic, and 3.4% reporting some other. Data collection included the Semi-Structured Assessment for the Genetics of Alcoholism (SSAGA), (Bucholz et al., 1994), neurophysiological and neuropsychological protocols, and collection of blood for DNA (Dick et al., 2023). In 2004, COGA began the prospective study of adolescents and young adults, targeting assessment of youth aged 12-22 from COGA families where at least one parent had been interviewed (Bucholz et al., 2017). These subjects were re-assessed every two years; currently, 89% of those individuals have 2+ interviews. We analyzed this prospective sample of adolescent and young adult offspring (N = 3,721).

### Substance use disorders (SUD)

Our measures of substance use disorders, both lifetime diagnosis and maximum severity, were derived from SSAGA interviews. We used the most severe occurrence of DSM-5 diagnosis for alcohol, cannabis, opioid, cocaine, and other SUD (stimulants or sedatives). We lacked DSM-5 diagnoses for tobacco/nicotine and used DSM-IV nicotine dependence instead.

For severity, we used the maximum reported DSM-5 lifetime symptom count of each substance, except nicotine. For nicotine/tobacco dependence, we indexed severity using a count of items from the Fagerstrom Test of Nicotine Dependence (FTND; Heatherton et al., 1991).

### Early life and adult socioeconomic status

We used data from SSAGA interviews of the prospective COGA participants and earlier SSAGA interviews completed by their parents to construct measures of early-life and adult SES. For parental education, we used the larger value (in years of education) reported by either the mother or father in cases where both were present. Education of the participants in the prospective study was the highest reported level of education (in years of education) at the last recorded observation. Household income for both parents and prospective participants was measured using a 9-item ordinal scale ranging from 1) $1-$9,999/year to 9) $150,000+/year. We converted the ordinal scale to a pseudo-continuous measure of income, using the midpoint for the first eight categories and the harmonic mean for the top category (von Hippel et al., 2016). Both parent and adult income were converted to 2024 dollars using the priceR (version 1.0.4, Condylios, 2026) package in R (version 4.5) to adjust for inflation. Descriptive statistics, missingness, and correlations between parents are presented in Supplemental Table 1.

### Change in socioeconomic status

We created an index of change in SES between SES-of-origin (e.g., early life SES) and achieved SES (e.g., adult SES). First, we created a composite index by adding education (1 = less than high school, 2 = high school or equivalent, 3 = some college, 4 = bachelor’s degree, 5 = graduate degree) and income (1 = bottom quintile to 5 = top quintile) for both childhood and adulthood (ranges of 2 - 10, for each). Next we took the difference between the adult SES composite and the childhood SES composite, resulting in an overall change score ranging from a possible-8 to 8. Scores greater than zero were indicative of upward mobility, scores less than zero were indicative of downward mobility, and scores of zero indicate no change between parents and offspring.

### Demographic and other covariates

To adjust for potential confounding, we included the age of participants at last observation, sex (male or female), and race-ethnicity (Non-Hispanic White, Black or African American, Hispanic or Latino/a/x, and other race-ethnicity) as covariates. Additionally, because COGA was ascertained based on family density of alcohol use disorders, there is detailed pedigree information on the affected status of multiple relatives. We used a previously constructed measure of family history density (FHD) for alcohol use disorder to estimate familial risk for SUD (Pandey et al., 2020; Rice et al., 1995).

### Analytic Strategy

Our analytic strategy was composed of several steps. First, we examined the contribution of early life SES to both diagnosis and severity for each of the SUD outcomes in the full sample. Second, among those participants old enough to have passed through the life course period in which typical educational milestones have been completed (those ≥ 25 years at age of last observation, N = 1,844) (Bureau of Labor Statistics, 2025), we compared the relative influence of early life and adult SES. For binary outcomes (e.g., diagnosis) we tested associations using logistic regression, applying robust standard errors to correct for clustering at the family level. For count measures (e.g., severity), we fit a series of negative binomial models to test the association between both early life and adult SES and SUD-severity. For each outcome, we present: 1) the estimates for the SES measures only, 2) models adjusted for age (at last observation), gender, and race-ethnicity, and 3) models adjusted for sociodemographic covariates and FHD. To correct for familial clustering, we applied cluster robust standard errors (Cameron et al., 2011; Zeileis, 2006). Lastly, we applied a false discovery rate of 5% to account for multiple testing (Benjamini and Hochberg, 1995).

## Results

### Descriptives

Table 1 displays the basic descriptives for the full, prospective cohort sample and participants who were at or over the age of 25 at their last observation. Lifetime prevalence and symptom severity follow a similar pattern across the full sample and age 25+ subset, with AUD, CUD, and TUD being the most prevalent in that order. The same pattern is observed for the aged 25+ subset, though the means/proportions are higher in this subgroup, likely reflective of the greater age in this sample and more time for these participants to have passed through the typical period of SUD onset. On average, parents and participants had completed slightly more education than a high school diploma in both the full sample and age 25+ subset. As expected, the mean reported years of education was greater in the age 25+ subset. Importantly, the full prospective sample and the age 25+ subset did not significantly differ across gender, race-ethnicity, parental education or income, or FHD.

**Table 1:** Demographic characteristics of the COGA Prospective Sample.

|  | Full sample<br>(N = 3,721) |  | Aged 25+<br>(N = 1,844) |  |
| --- | --- | --- | --- | --- |
|  | <u>Mean/N</u> | <u>SD/%</u> | <u>Mean/N</u> | <u>SD/%</u> |
| Age at last observation (years) | 25.07 | 6.90 | 31.14 | 3.44 |
| Female | 1,903 | 51.22% | 1,007 | 54.61% |
| Non-Hispanic White | 2,233 | 60.12% | 1,095 | 59.38% |
| Black or African American | 933 | 25.12% | 469 | 25.43% |
| Hispanic or Latino/a/x | 423 | 11.39% | 226 | 12.26% |
| Other race or ethnicity | 125 | 3.37% | 54 | 2.93% |
| Family history density of alcohol use disorder | 0.39 | 0.16 | 0.40 | 0.16 |
| Parental education (years) | 13.47 | 2.19 | 13.00 | 2.16 |
| Adult education (years) | 12.99 | 2.51 | 14.00 | 2.01 |
| Parental income (household)* | 93,879 | 78,286 | 95,707 | 78,703 |
| Adult income (household)* | 108,305 | 77,512 | 119,240 | 75,165 |
| Alcohol use disorder (lifetime) | 1,441 | 38.73% | 950 | 51.52% |
| Cannabis use disorder (lifetime) | 1,232 | 33.11% | 728 | 39.48% |
| Tobacco use disorder (lifetime) | 911 | 24.48% | 578 | 31.34% |
| Cocaine use disorder (lifetime) | 179 | 4.81% | 131 | 7.10% |
| Opioid use disorder (lifetime) | 192 | 5.16% | 133 | 7.21% |
| Other substance use disorder (lifetime) | 210 | 5.64% | 147 | 7.97% |
| Alcohol use disorder (severity) | 1.77 | 2.41 | 2.44 | 2.69 |
| Cannabis use disorder (severity) | 1.76 | 2.77 | 2.17 | 3.02 |
| Tobacco use disorder (severity) | 1.29 | 2.34 | 1.67 | 2.60 |
| Cocaine use disorder (severity) | 0.30 | 1.47 | 0.46 | 1.81 |
| Opioid use disorder (severity) | 0.36 | 1.67 | 0.51 | 1.98 |
| Other substance use disorder (severity) | 0.37 | 1.62 | 0.52 | 1.90 |

\* value converted to 2024 dollars

Correlations between SES measures and SUDs are presented in Figure 1. There were modest correlations between education and income in the parents (r = 0.47) and participants (r = 0.46). Parental income was moderately correlated with participant income in adulthood (r = 0.42) but the relationship between parental education and highest level of education achieved by the participants was weaker (r = 0.28). There were moderate to strong correlations between the SUDs (r = 0.50-0.79). Parental and adulthood SES variables had weak negative correlations with SUD diagnoses in adulthood. The one exception was a weak positive correlation between AUD and highest level of education attained in adulthood (r = 0.22).

**Figure 1:**
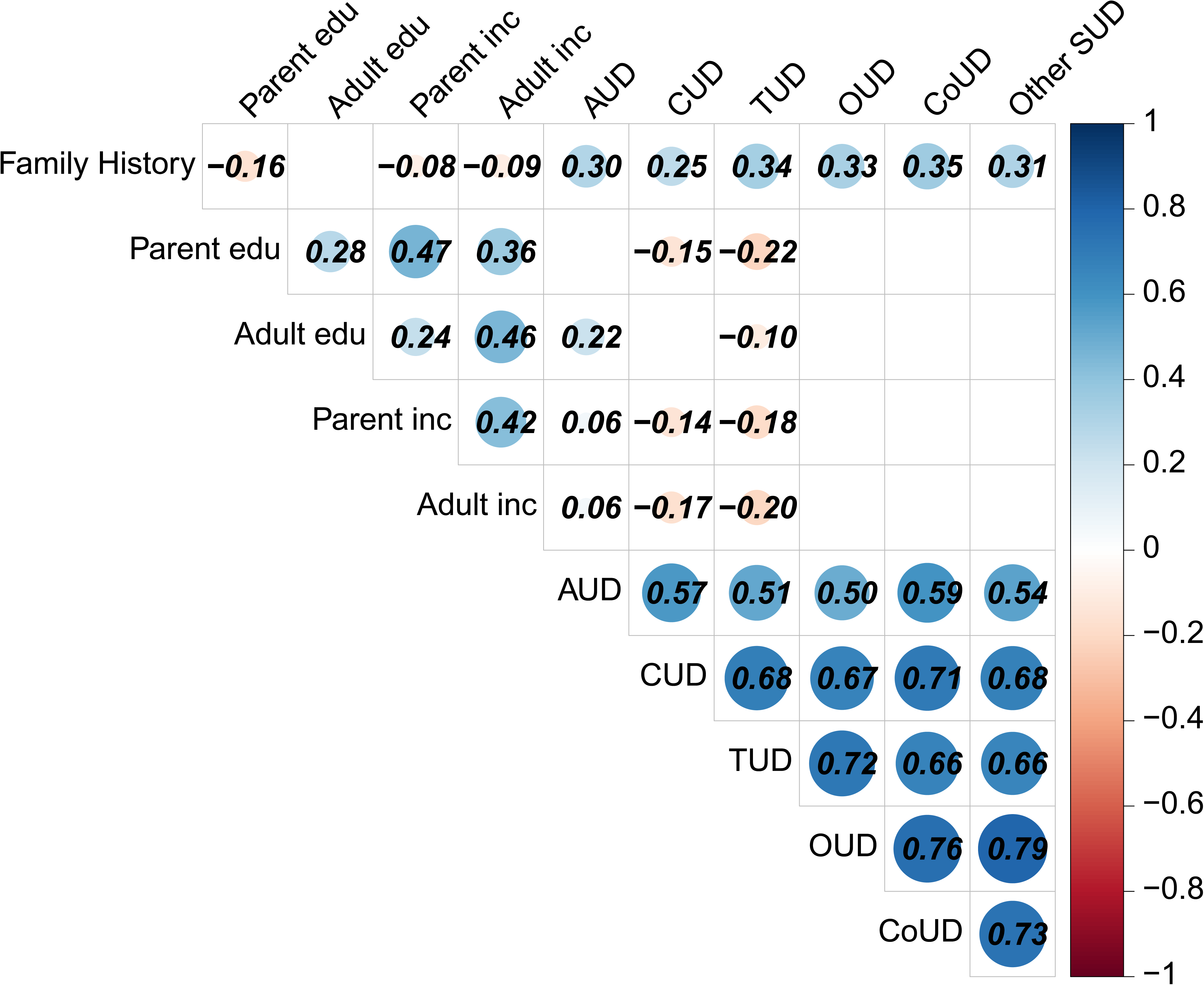
Correlations Between SES and SUD Outcomes in COGA Prospective Participants (N = 3,721). Estimates for Pearson’s (continuous-continuous), polyserial (continuous-binary), and tetrachoric (binary-binary) correlations. Blank cells indicate estimates with *p* >.05. Color scale provides the range of correlation values. AUD = alcohol use disorder, TUD = tobacco use disorder, CUD = cannabis use disorder, CoUD = cocaine use disorder, OUD = opioid use disorder, Other SUD = other substance use disorder.

### Early Life SES and SUD Outcomes in Adulthood

Figure 2 displays the associations between early life SES and SUD outcomes across a series of models with increasing levels of covariate control. For the unadjusted results (SES measures only), the associations between early life SES were strongest with TUD outcomes, such that higher levels of parental education and income were associated with decreased risk of TUD diagnosis (OR_P-EDU_ = 0.72, 95% CI = 0.65, 0.79; OR_P-INC_ = 0.85, 95% CI = 0.76, 0.95) and severity (IRR_P-EDU_ = 0.76, 95% CI = 0.70, 0.82; IRR_P-INC_ = 0.84, 95% CI = 0.77, 0.92). The associations with lifetime TUD diagnosis (OR_P-EDU_ = 0.73, 95% CI = 0.66, 0.82 OR_P-INC_ = 0.80, 95% CI = 0.71, 0.90) and TUD severity (IRR_P-EDU_ = 0.76, 95% CI = 0.70, 0.82; IRR_P-INC_ = 0.84, 95% CI = 0.77, 0.92) remained after accounting for both demographics and family history.

**Figure 2:**
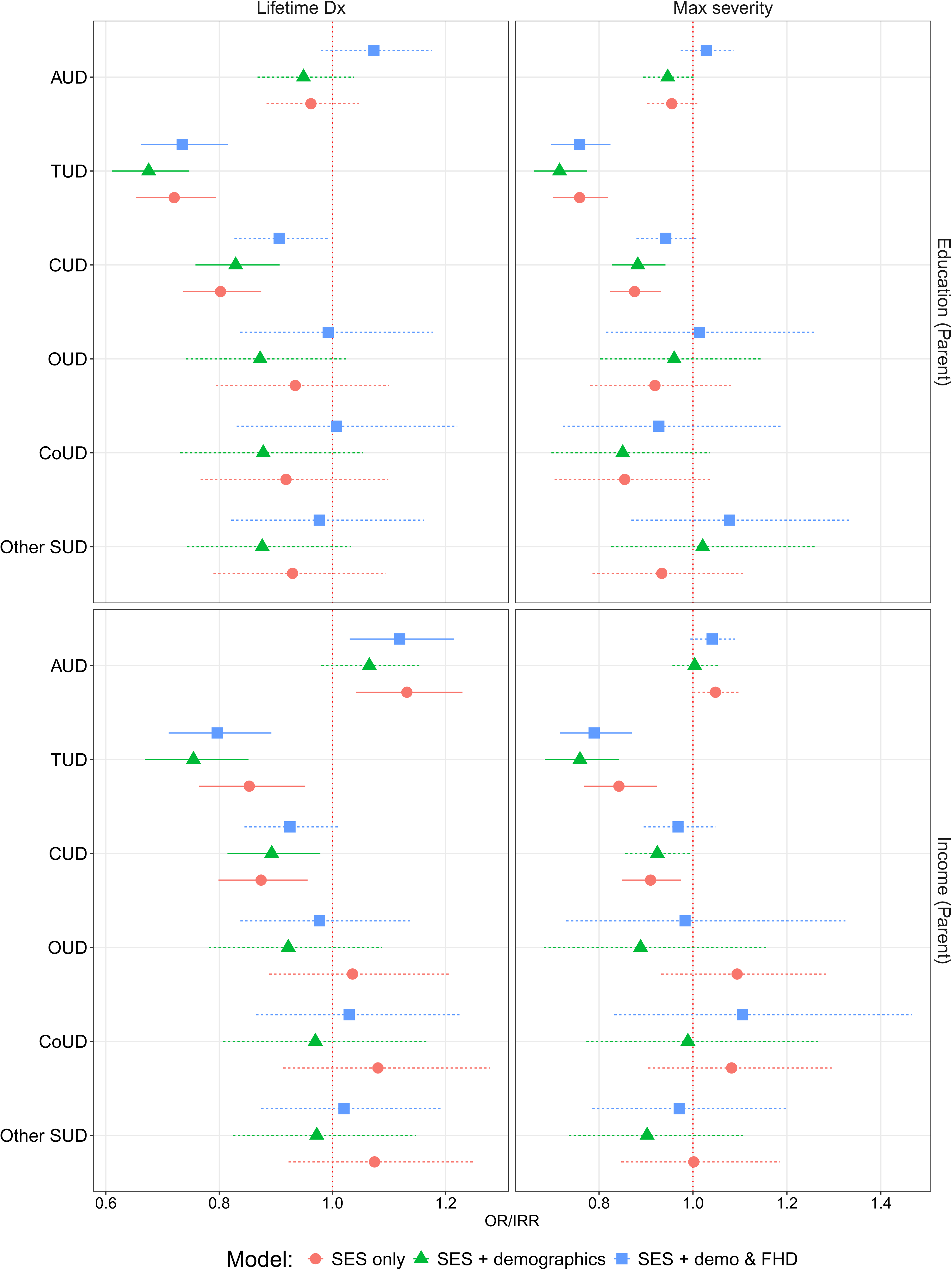
Estimates for Early Life SES with SUD Outcomes in COGA Prospective Participants (N = 3,721). Point estimates (and 95% confidence intervals) for logistic regression models focusing on lifetime diagnosis (odds ratios, or OR) and negative binomial models focusing on symptom severity (incident rate ratios, or IRR). Red lines indicate models with only parental education and income. Green lines indicate models with parental education, income, age, gender, and race-ethnicity. Blue lines indicate models with parental education, income, age, gender, race-ethnicity, and family history density of alcohol use disorder. AUD = alcohol use disorder, TUD = tobacco use disorder, CUD = cannabis use disorder, CoUD = cocaine use disorder, OUD = opioid use disorder, Other SUD = other substance use disorder.

Early life SES was also significantly negatively associated with CUD diagnosis (OR_P-EDU_ = 0.80, 95% CI = 0.74, 0.87; OR_P-INC_ = 0.87, 95% CI = 0.80, 0.96) and severity (IRR_P-EDU_ = 0.88, 95% CI = 0.82, 0.93; IRR_P-INC_ = 0.91, 95% CI = 0.85, 0.97). The associations with lifetime CUD diagnosis (OR_P-EDU_ = 0.83, 95% CI = 0.76, 0.91; OR_P-INC_ = 0.89, 95% CI = 0.81, 0.98) and severity (IRR_P-EDU_ = 0.88, 95% CI = 0.83, 0.94) remained once demographic covariates were added to the models for all but the association between CUD severity and parental income. Including family history density as a covariate further attenuated results and none of the CUD associations remained significant after correcting for multiple testing.

Finally, there was a significant positive association between parental income and AUD diagnosis in the SES-only models (OR_P-INC_ = 1.13, 95% CI = 1.04, 1.23). Covarying for demographics and family history density in the models for lifetime AUD diagnosis did not alter the association with parental income (OR_P-INC_ = 1.12, 95% CI = 1.03, 1.21). There were no significant associations between any of the early life SES measures and OUD, CoUD, or other SUDs in any of the models. Finally, we ran all of the above models in the subset aged ≥ 25. The results were virtually identical when limiting to the age 25+ subset used in subsequent analyses (full results are presented in Supplementary Tables 2-5).

### Relative Contributions of Early Life SES and Adult SES to SUD Outcomes in Adulthood

Figure 3 displays the results from the series of models that examined the associations between early life and adult SES with SUD outcomes among COGA participants that had provided data at or beyond the age of 25. In contrast to the early life SES effects, years of education in adulthood showed significant negative associations across all SUD outcomes. Conditional on early life SES (parent education and income) and current household income, adult education was associated with reduced risk for diagnosis and lower severity in alcohol (OR_A-EDU_ = 0.77, 95% CI = 0.66, 0.89; IRR_A-EDU_ = 0.80, 95% CI = 0.74, 0.87), tobacco (OR_A-EDU_ = 0.39, 95% CI = 0.32, 0.46; IRR_A-EDU_ = 0.47, 95% CI = 0.41, 0.53), cannabis (OR_A-EDU_ = 0.42, 95% CI = 0.36, 0.50; IRR_A-EDU_ = 0.62, 95% CI = 0.56, 0.69), opioid (OR_A-EDU_ = 0.43, 95% CI = 0.32, 0.57; IRR_A-EDU_ = 0.39, 95% CI = 0.28, 0.56), cocaine (OR_A-EDU_ = 0.46, 95% CI = 0.34, 0.61; IRR_A-EDU_ = 0.47, 95% CI = 0.35, 0.64), and other substance use disorders (OR_A-EDU_ = 0.42, 95% CI = 0.32, 0.55; IRR_A-EDU_ = 0.37, 95% CI = 0.29, 0.47). The addition of demographic covariates and family history density to these models did not result in a significant attenuation of the effect sizes.

**Figure 3:**
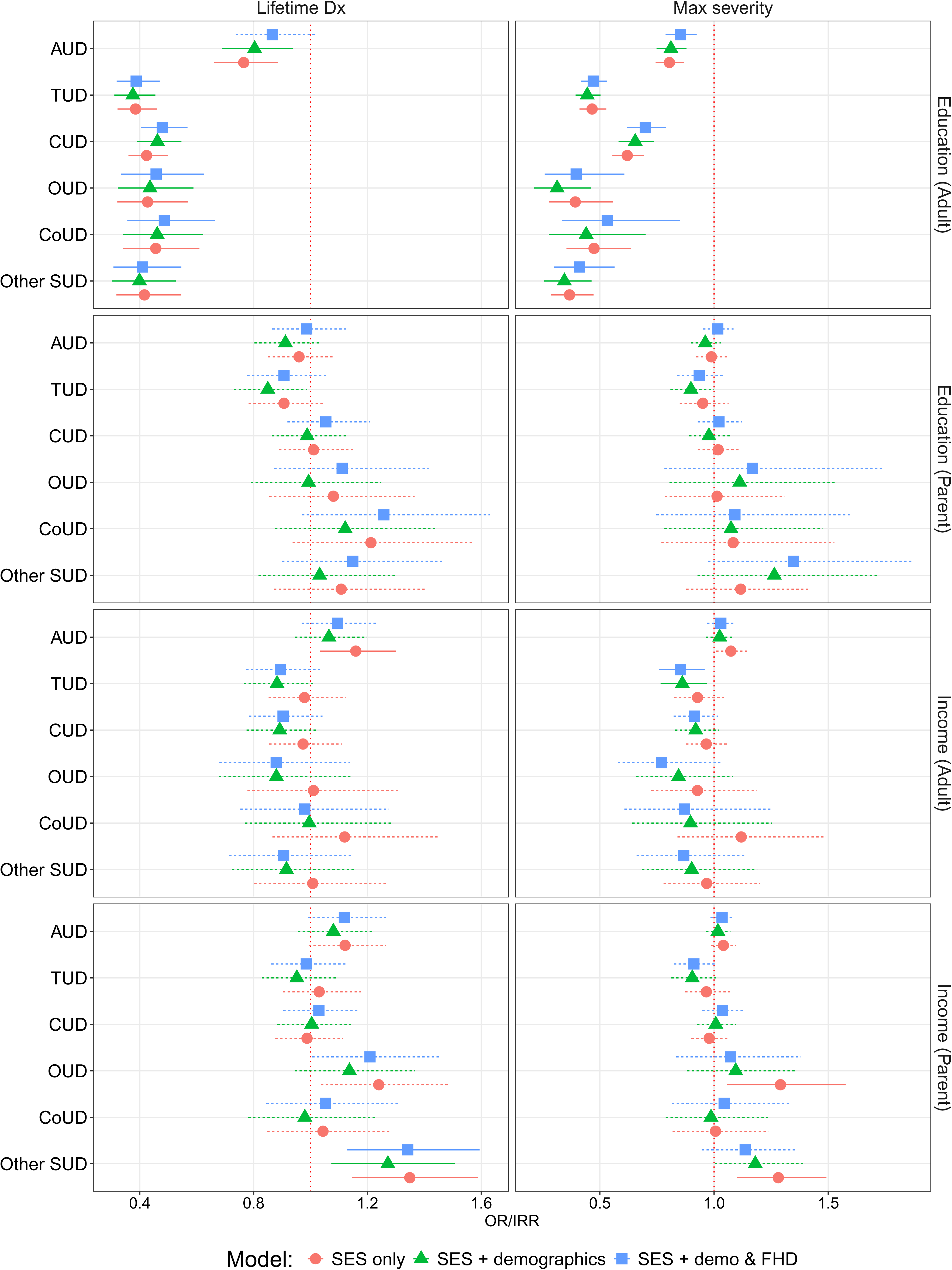
Estimates for Early Life & Adult SES with SUD in COGA Prospective Participants age 25 and older (N = 1,844). Point estimates (and 95% confidence intervals) for logistic regression models focusing on lifetime diagnosis (odds ratios, or OR) and negative binomial models focusing on symptom severity (incident rate ratios, or IRR). Red lines indicate models with only adult education, adult income, parental education, and parental income. Green lines indicate models that also include, age, gender, and race-ethnicity. Blue lines indicate models that also include family history density of alcohol use disorder. AUD = alcohol use disorder, TUD = tobacco use disorder, CUD = cannabis use disorder, CoUD = cocaine use disorder, OUD = opioid use disorder, Other SUD = other substance use disorder.

Income in adulthood was significantly associated with an AUD diagnosis in the unadjusted models (OR_A-INC_ = 1.16, 95% CI = 1.03, 1.30), such that higher income levels were associated with increased risk of having an AUD. This association was no longer significant after adding demographic covariates and family history density to the model. Income in adulthood was not significantly associated with any other SUD outcomes. For most of the SUD outcomes considered, early life SES measures attenuated with the addition of adult measures of SES. The one exception to this trend was the significant positive association between parental income and diagnosis of other SUDs, which was significant even after adjusting for demographics and family history (OR_P-INC_ = 1.34, 95% CI = 1.13, 1.59). The full results are presented in Supplementary Tables 6-7.

Lastly, as a set of descriptive analyses, we explored mean SUD severity across the change SES between parent and participants. The descriptive results are depicted in Figure 4. These exploratory results provide evidence for two important patterns: 1) upwardly mobile individuals had lower risk while downwardly mobile individuals had highest risk for TUD, CUD, OUD, and other SUDs, suggesting processes of both social selection and social causation may be at play for these substances, and 2) the relative change in SES may be as relevant as absolute levels of SES in SUD risk.

**Figure 4:**
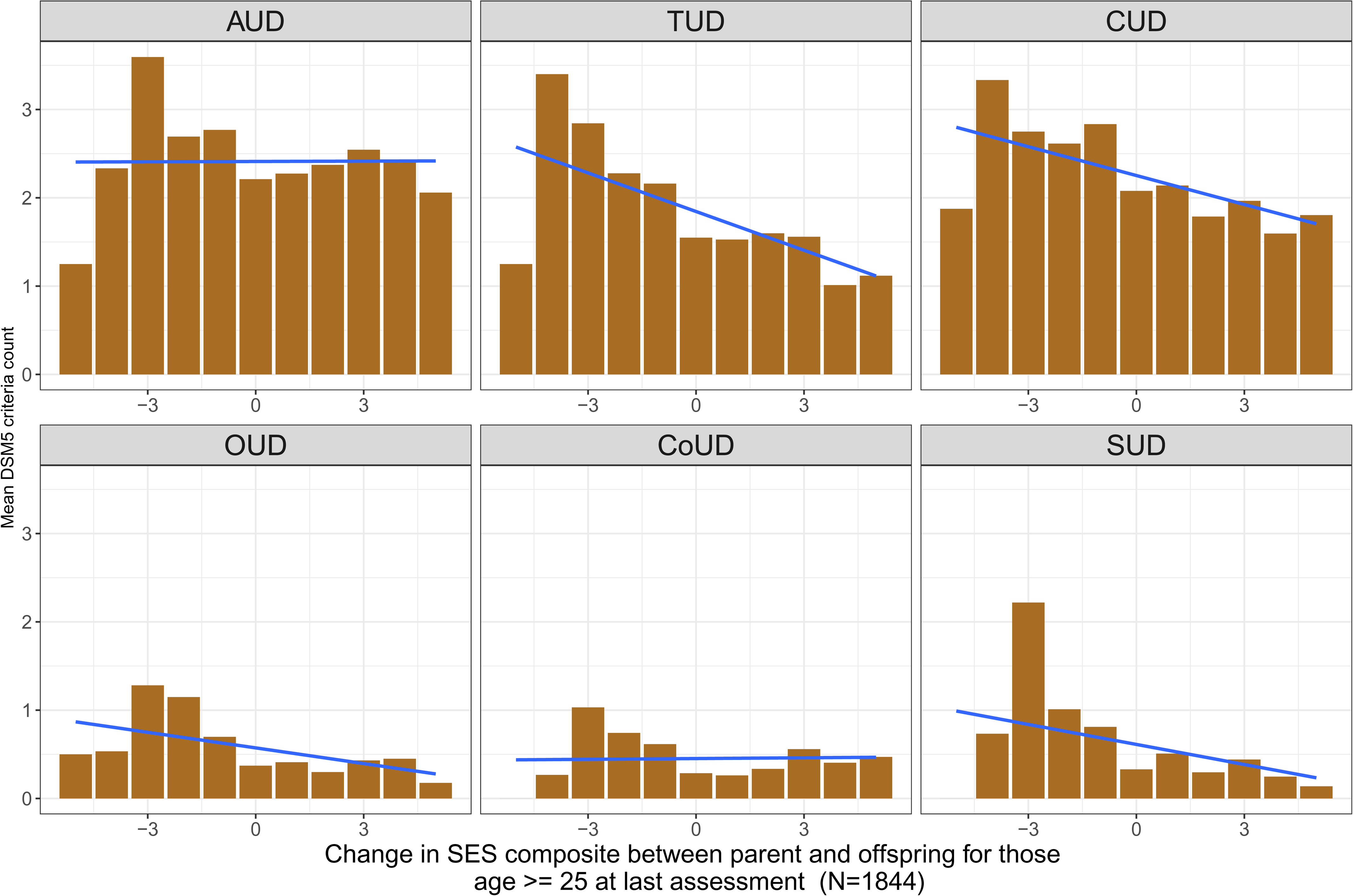
Changes in SES and SUD Severity in COGA Prospective Participants age 25 and older (N = 1,844). Mean symptoms counts (and best fit line) across difference in a composite index of education (1 = less than high school to 5 = graduate degree) and income (1 = bottom quintile to 5 = top quintile) children and parents (ranging from a possible-10 to 10) change scores for each substance use disorder. Scores greater than zero indicate upward mobility and negative values indicate downward mobility. AUD = alcohol use disorder, TUD = tobacco use disorder, CUD = cannabis use disorder, CoUD = cocaine use disorder, OUD = opioid use disorder, Other SUD = other substance use disorder.

## Discussion

Socioeconomic inequalities can have profound effects on patterns of morbidity and mortality within the broader population (Braveman et al., 2011). Research over the past 50 years has consistently found strong gradients in health across SES. In the current study, we explored the relationship between both early life and adult SES with SUD prevalence and severity in COGA, a family-based study enriched for SUDs. Briefly, we found that early life SES, particularly parental education, is negatively associated with subsequent diagnosis and severity of several SUD (e.g., cannabis, nicotine). When examining the joint effect of early life and adult SES, higher adult education, alone, was associated with a lower risk of both diagnosis and severity for all of the SUDs considered. Finally, descriptive explorations of change in SES (difference from that of parents) revealed those that were upwardly mobile had the lowest severity and conversely those that were downwardly mobile had the greatest severity across most SUD.

Early life SES (both parent income and education) was relevant for alcohol, cannabis, and tobacco use disorders, with some variation in the direction of effect. The positive association between parental income and AUD diagnosis runs counter to prior results from population-based samples, which found early life SES was negatively associated with young adult alcohol problems (Kendler et al., 2014; Lee et al., 2018). However, there is considerable variation in directions of effect across studies and type of alcohol use outcomes considered (e.g., frequency, quantity, or problems) and the positive association between income and normative consumption instead of problems is a consistent finding (Kendler et al., 2014; Lee et al., 2018; Patrick et al., 2012). Our unexpected findings may also reflect the clinically-ascertained nature of the COGA sample, and the potentially non-linear effects of parental income for youth who have heightened genetic and environmental vulnerabilities to developing an SUD. For both tobacco and cannabis, parental education and income were associated with both lifetime diagnosis and SUD severity. For tobacco, results mirror prior findings that show early life SES is negatively associated with adult smoking status (Patrick et al., 2012) and that conditions related to early life SES, such as neighborhood stability and disadvantage, are associated with adult tobacco use disorder symptoms (Lee et al., 2018). There is also some evidence that early life SES is negatively related to future cannabis use disorder (Daniel et al., 2009; Lee et al., 2018). Overall, these results broadly support the idea that early life SES is an important life course exposure that can have consequences for future SUD risk, with variability across substances.

When exploring the joint effect of early life and adult SES, adult educational attainment emerged as the most relevant SES facet for both diagnosis and severity across all SUD types. The negative association between educational attainment in adulthood and SUD diagnosis and severity mimics previous evidence (Fothergill et al., 2008; Green et al., 2012; Kessler et al., 1995; Martin et al., 2015). Prior research also supports independent effects of education and income on health outcomes (Backlund et al., 1999; Braveman et al., 2005; Cundiff et al., 2015; Herd et al., 2007). However, ours is the first study to parse the effects of these SES domains for SUD diagnosis and severity across so many forms of SUD, simultaneously. Our finding that the inclusion of SES in adulthood (specifically educational attainment) diminishes the effects of childhood/parental SES has mixed support in the literature. Some studies have reported a unique effect of childhood/parental SES while controlling for current SES (Poulton et al., 2002). Other studies mirror our findings that SES in adulthood is more strongly associated with SUDs and other health outcomes than SES in childhood (Lynch et al., 1994). Given the relationship between early life and adult SES, it is plausible that any impact of early life SES (and the adversities associated with it) is mediated through its impact on adult SES (Suglia et al., 2022).

Finally, the exploratory analyses examining changes in SES from childhood to adulthood revealed that change in SES, not just one’s absolute starting position, was an important indicator of risk for tobacco, cannabis, opioid, and other substance use disorders; but not for alcohol or cocaine. Upward socioeconomic mobility from childhood to adulthood was associated with fewer symptoms for TUD, CUD, OUD, and other SUDs. Previous research has found positive associations between upward mobility and health outcomes (Barakat and Konstantinidis, 2023; Cundiff et al., 2017; Dennison, 2018), including mental health and substance use outcomes However, these positive effects may vary by sex and race/ethnicity (Hudson et al., 2026; Zang and Tian, 2025). With regard to AUD and cocaine use disorder (CoUD), we found no association between change in SES and severity or likelihood of disorder. This mirrors findings from prior analyses that found that upward mobility did not mitigate the effects of low childhood SES on adult health, including both alcohol and tobacco dependence (Poulton et al., 2002). Some research has shown that affluence is associated with increased substance use, especially alcohol (Martin, 2019; Patrick et al., 2012), which may explain the null associations between change in SES and AUD in the current study. However, there is also evidence that frequency, quantity, and problems have unique patterns of associations with SES (Kendler et al., 2014), Future analyses should explore whether the positive associations with AUD remain after accounting for drinking frequency and quantity.

This research has several important limitations. First, we examined lifetime diagnosis and maximum symptom severity and did not consider the timing of SUD onset, which may be relevant for teasing apart processes of social selection vs. social causation. Second, we focused on two key components of SES: income and education. We did not explore other possible aspects that may be relevant including occupation, wealth, or neighborhood-related aspects of SES. Finally, we included family history density of AUD as a way of assessing familial risk, an important consideration for SUD. Other measures of genetic risk, such as polygenic scores, may be less confounded by the environmental aspects of familial risk and can be applied in samples such as COGA (which contain measured genetic data). However, current polygenic scores in psychiatric genetics are still plagued by an overreliance on samples of European descent and perform relatively worse in other populations (Ding et al., 2023; Mills and Rahal, 2019; Peterson et al., 2019). FHD is still more predictive than polygenic scores when leveraging the diverse participants available in COGA (Hujoel et al., 2022), though both FHD and polygenic scores may capture unique variance in SUDs (Lai et al., 2022; Mars et al., 2022).

Socioeconomic inequalities have implications for variation in SUDs. Our findings yield important understanding of the etiology of substance use disorders in a multi-faceted socioenvironmental context, with implications for public health prevention across the lifespan. Both early life socioeconomic advantage and greater adult education were associated with reduced risk of lifetime diagnosis and lower severity across multiple SUDs, even after accounting for early life SES, familial risk, and sociodemographic covariates. Those that were upwardly mobile had lowest severity in SUDs while the downwardly mobile exhibited the opposite pattern, suggesting processes of both social causation and social drift. In total, these results support the notion that socioeconomic inequalities are relevant for SUDs, similar to other health conditions and that a life course framework is important for understanding of the role of SES in SUD development. While treatment efforts for SUDs often occur at the individual level, policies that seek to alleviate the negative conditions associated with economic inequality are also necessary to help prevent and reduce the impact of these chronic conditions.

## Data Availability

COGA data available through dbGaP (Study Accession: phs000763.v1.p1).

## Acknowledgements

This work was funded by the National Institute of Drug Abuse (R01DA061850). This publication does not represent the views of the National Institutes of Health, or the United States Government. Dr. Barr had full access to all the data in the study and takes responsibility for the integrity of the data and the accuracy of the data analysis. The funders had no role in the design and conduct of the study; collection, management, analysis, and interpretation of the data; preparation of the manuscript; and decision to submit the manuscript for publication. The COGA Publication Committee reviewed and approved the manuscript.

The Collaborative Study on the Genetics of Alcoholism (COGA), Principal Investigators B. Porjesz, V. Hesselbrock, A. Agrawal; Scientific Director, A. Agrawal; Translational Director, D. Dick, includes nine different centers: University of Connecticut (V. Hesselbrock); Indiana University (H.J. Edenberg, T. Foroud, Y. Liu, M.H. Plawecki); University of Iowa Carver College of Medicine (A. Andersen S. Kuperman); SUNY Downstate Health Sciences University (B. Porjesz, J. Meyers); Washington University in St. Louis (L. Bierut, A. Agrawal, S. Hartz); University of California at San Diego (M. Schuckit); Rutgers University (D. Dick, R. Hart, J. Salvatore, J. Tischfield); The Children’s Hospital of Philadelphia, University of Pennsylvania (L. Almasy); Icahn School of Medicine at Mount Sinai (A. Goate, P. Slesinger); and Howard University (D. Scott). Other COGA collaborators include: M. Hesselbrock, K. Manning (University of Connecticut); D. Lai, J. Nurnberger Jr., L. Wetherill, A. Miller, X. Xuei, (Indiana University); J. Kramer (University of Iowa), G. Chan (University of Iowa; University of Connecticut); C. Kamarajan, A. Pandey, D.B. Chorlian, P. Barr, S. Kinreich, G. Pandey, Z. Neale, C. Chatzinakos, J. Zhang, S. Saenz deViteri, A. Bingly (SUNY Downstate); G. Pathak (Icahn School of Medicine at Mount Sinai); A. Anokhin, K. Bucholz, F. Dong, A. Hatoum, E. Johnson, J. Rice, S. Saccone (Washington University); F. Aliev, Z. Pang, S. Kuo, S. Brislin, (Rutgers University). We continue to be inspired by our memories of Henri Begleiter and Theodore Reich, founding PI and Co-PI of COGA, and also owe a debt of gratitude to other past organizers of COGA, including Ting-Kai Li, P. Michael Conneally, Raymond Crowe, and Wendy Reich, for their critical contributions. This national collaborative study is supported by NIH Grant U10AA008401 from the National Institute on Alcohol Abuse and Alcoholism (NIAAA) and the National Institute on Drug Abuse (NIDA).

